# AI-BASED MRI MONITORING IN MULTIPLE SCLEROSIS: REAL-WORLD CLINICAL VALIDATION

**DOI:** 10.1101/2023.08.14.23293959

**Authors:** Michael Barnett, Dongang Wang, Heidi Beadnall, Antje Bischof, David Brunacci, Helmut Butzkueven, J William L Brown, Mariano Cabezas, Tilak Das, Tej Dugal, Daniel Guilfoyle, Alexander Klistorner, Stephen Krieger, Kain Kyle, Linda Ly, Lynette Masters, Andy Shieh, Zihao Tang, Anneke van der Walt, Kayla Ward, Heinz Wiendl, Geng Zhan, Robert Zivadinov, Yael Barnett, Chenyu Wang

## Abstract

Modern management of MS targets No Evidence of Disease Activity (NEDA): no clinical relapses, no magnetic resonance imaging (MRI) disease activity and no disability worsening. While MRI is the principal tool available to neurologists for monitoring clinically silent MS disease activity and, where appropriate, escalating treatment, standard radiology reports are qualitative and may be insensitive to the development of new or enlarging lesions. Existing quantitative neuroimaging tools lack adequate clinical validation. In 397 multi-center MRI scan pairs acquired in routine practice, we demonstrate superior case-level sensitivity of a clinically integrated AI-based tool over standard radiology reports (93.3% vs 58.3%), relative to a consensus ground truth, with minimal loss of specificity. We also demonstrate equivalence of the AI-tool with a core clinical trial imaging lab for lesion activity and quantitative brain volumetric measures, including percentage brain volume loss (PBVC), an accepted biomarker of neurodegeneration in MS (mean PBVC -0.32% vs -0.36% respectively), whereas even severe atrophy (>0.8% loss) was not appreciated in radiology reports. Finally, the AI-tool additionally embeds a clinically meaningful, experiential comparator that returns a relevant MS patient centile for lesion burden, revealing, in our cohort, inconsistencies in qualitative descriptors used in radiology reports. AI-based image quantitation enhances the accuracy of, and value-adds to, qualitative radiology reporting. Scaled deployment of these tools will open a path to precision management for patients with MS.

## INTRODUCTION

Multiple sclerosis (MS) is the most common inflammatory demyelinating and neurodegenerative condition of the central nervous system (CNS), afflicting some 2.8 million persons globally [1]. Characterised by both focal lesions and by more diffuse neurodegeneration in the brain and spinal cord, MS results in significant physical and cognitive disability and, in many cases, premature withdrawal from the workforce.

Highly effective disease modifying therapy (DMT) dramatically reduces the risk of relapse associated worsening (RAW), but has little effect on progression independent of relapse activity (PIRA), the principal driver of increasing disability in patients with established, treated disease [2,3]. Inflammatory activity, the pathological substrate for RAW, and response to DMT are monitored by regular clinical assessment and repeated magnetic resonance imaging (MRI), usually on an annual basis [4]. MRI is also the most important tool for neurologists to assess disease activity that does not manifest with overt clinical change, but potentially injures vast numbers of axons and disrupting complex integrated brain networks. Specifically, the development of new or enlarging hyperintensities on FLAIR MRI and/or new contrast enhancing lesions (CELs) on T1-w MRI generally suggests inadequate suppression of inflammatory activity and may prompt the clinician to change the patient’s DMT [4]. PIRA is more difficult to characterise by MRI, but at the group level disability worsening correlates well with whole brain volume change [5,6]. In the absence of available DMTs that specifically target neurodegeneration, MRI evidence of accelerated brain atrophy, which at the individual level can be confounded by biological, disease- and treatment-related fluctuations, is generally not used in isolation to drive treatment change. However, there is broad agreement that the modern management of MS should target ‘NEDA-3’, or No Evidence of Disease Activity (no clinical relapses, no MRI activity, no disability worsening) [7].

The radiologist therefore plays a critical role, not only in the diagnosis of MS, but in the monitoring of the disease and its response to DMT. Traditionally, detailed slice-by-slice examination of current and prior study FLAIR images is required to accurately exclude the development of new or enlarging lesions, a painstaking process that has become increasingly burdensome with the advent of 3D imaging, which generates up to 300 slices in a single volume. Lack of current and prior 3D FLAIR volume co-registration in many picture archiving and communications systems (PACS) can also hamper the accurate detection of small new lesions or minor lesion enlargement, particularly when concentric, between studies. While the volume of new (or enlarging) lesions may impact treatment strategy, this is not measured or reported in routine clinical radiology practice. An estimation of the severity of the overall FLAIR lesion burden, which provides prognostic information, is also dependent on the experience of the reporting radiologist and can only be semi-quantitatively assessed. Severe brain volume loss (BVL) versus age-matched healthy controls, which may also be of prognostic significance, can be detected by experienced radiologists with visual inspection but cannot be accurately quantitated without additional tools, which are generally confined to research settings. Moderate changes (of the magnitude expected in many patients with MS) are difficult, if not impossible, to recognise by visual inspection alone [8,9]. Similarly, longitudinal change in brain volume during the typical 12-month interval between MRI scans is usually small and not detectable by visual inspection. While short-term changes in brain volume are difficult to interpret in individual patients, a consistent adverse trajectory over multiple clinical epochs or more severe brain atrophy (>0.8% percent BVL per annum) over a single epoch, may influence or support clinical decisions to escalate or switch DMT [4].

Recognition that clinical radiology reports for patients with MS can be enhanced by quantitative information has been accompanied, in the last 5 years, by the development of artificial intelligence (AI) algorithms for medical imaging that can automate both the detection and segmentation of the brain, brain substructures and different types of brain pathology, including MS lesions [10-13]. While there are a small number of existing commercial (regulatory-approved) image analysis tools that have been designed to assist radiologists and clinicians who treat patients with MS, thorough real-world clinical validation is limited [14]. Here, we report a comprehensive clinical evaluation of iQ-Solutions™ (MS Report), hereafter referred to as iQ-MS, in a large cohort of MS scan pairs that were independently reported in clinical practice by expert radiologists; and, separately, were quantitatively and blindly assessed by trained neuroimaging analysts in a core reading imaging laboratory using standard procedures (SOPs) used in regulatory trials. Specifically, we hypothesized that the AI tool would more sensitively and accurately detect MRI evidence of disease activity compared with conventional radiology reports; and produce cross sectional and longitudinal brain volumetric measurements comparable with those generated by conventional imaging tools implemented by the core lab.

### Core Technology

iQ-Solutions™ analyses brain MRI scans in Digital Imaging and Communications in Medicine (DICOM) format using a collection of AI algorithms based on deep neural network technology, and was developed using more than 8500 brain scans that had been expertly annotated by trained neuroimaging analysts. iQ-Solutions™ produces an MS-specific report that includes cross-sectional and longitudinal whole brain, brain substructure and lesion metrics relevant to the condition (Table 1). The AI tool returns visualizations of relevant segmentations to the PACS for radiologist review (Figure 1).

**Table 1.** iQ-Solutions™ MS Report output: metrics^#^.

| Cross-sectional metrics | Longitudinal changes from previous scan |
| --- | --- |
| FLAIR lesion number and volume | Longitudinal protocol warnings (T1 and FLAIR)<sup>§</sup> |
| FLAIR lesion volume patient* centile | New FLAIR lesion number and volume |
| T1-w contrast enhancing lesion number and volume | Enlarging FLAIR lesion number and volume |
| Normalized whole brain volume | Annualized percentage brain volume change |
| Normalized whole brain volume centile*,** |  |
| Normalized thalamus volume |  |
| Normalized thalamus volume HC centile** |  |

<sup>#</sup> iQ-MS experiential MS reference dataset only available in the tool's research mode
\*compared with iQ-MS underlying MS reference dataset
<sup>§</sup> provided in research mode when scanner/protocol inconsistencies detected (supplementary data)
\*\*compared with iQ-MS underlying healthy control dataset

**Figure 1.**
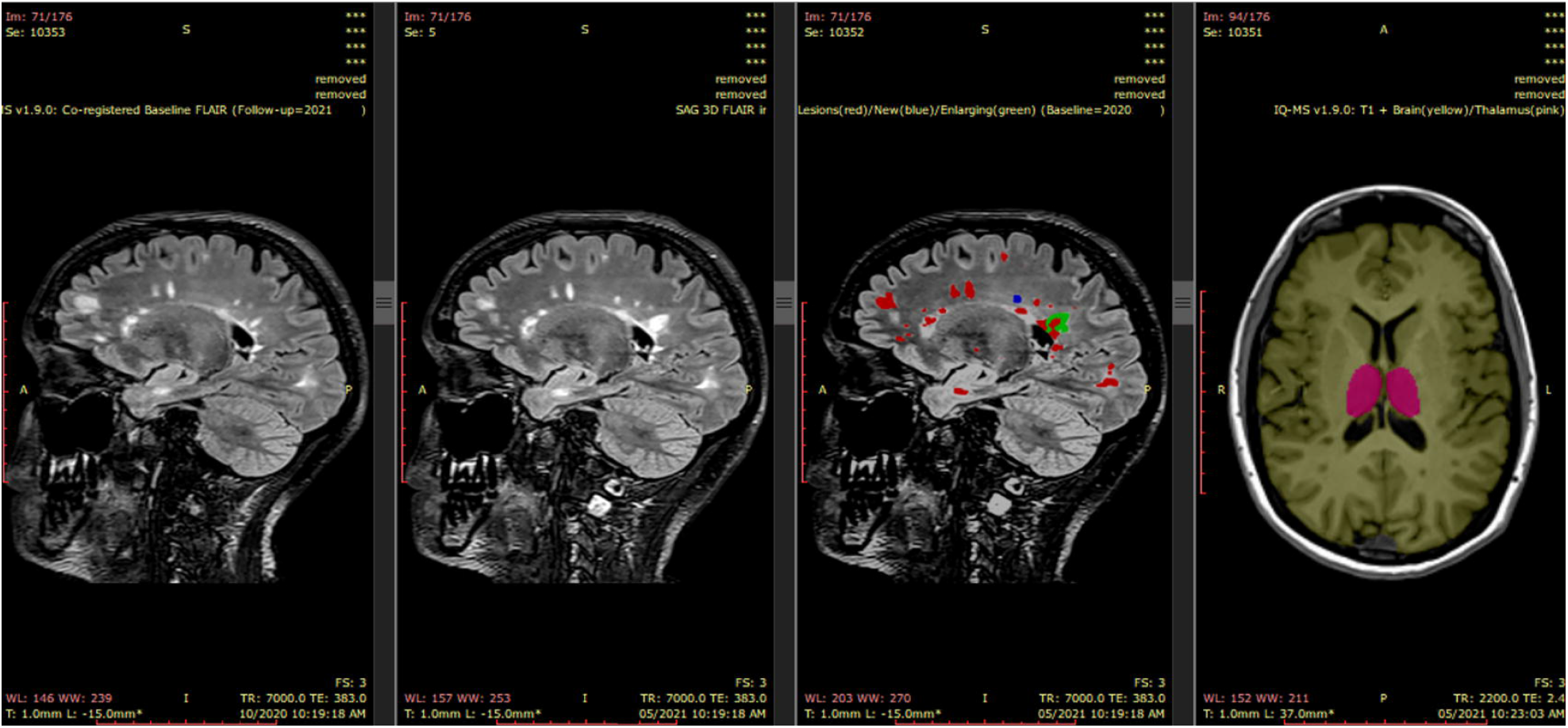
iQ-Solutions PACS integration iQ-MS automatically returns a co-registered baseline (prior study) 3D FLAIR series together with a lesion-annotated 3D FLAIR, here showing a case with both new (blue) and enlarging (green) lesions. A 3D-T1 series is also returned with both lesion-inpainted whole brain (green) and thalamus (pink) annotations.

For any analysis to proceed, images are automatically quality checked to ensure that pre-contrast 3D-T1 and 3D FLAIR sequences, each containing ≥30 slices with a thickness of ≤3mm, are available. All cross-sectional segmentation algorithms (Table 1) were developed with 3D-UNet [15] as the core network for extracting image features, followed by a solitary convolutional layer as the prediction head. Cross-validation was conducted through comparison (based on case-wise and voxel-wise DICE scores) with ground-truth masks produced by trained neuroimaging analysts. Similarly, lesion activity between timepoints (namely, the development of new and enlarging lesions) is measured by iQ-Solutions using an algorithm based on a modified 3D-Unet and trained with manually annotated 3D-FLAIR images, as described previously [16]. iQ-MS reports enlarging lesions as new lesional voxels that are connected to an existing lesion (on the prior study) within its 26-voxel neighborhood.

For brain and substructure volumetric analyses, a lesion-inpainting model, LG-Net, was applied to 3DT1 images to ameliorate segmentation bias generated by the presence of MS lesions, as previously described [17]. For longitudinal brain and brain substructure volumetric change, iQ-Solutions performs a number of checks for image consistency between the two scan timepoints (supplementary data). Longitudinal metrics are reported, but returned to the user with a protocol inconsistency warning. Longitudinal whole brain volume change is measured by iQ-MS with the integrated DeepBVC algorithm [18]. Automated estimation of substructure (whole grey matter, thalamus) volume change is produced by a combination of AI-based segmentation and the application of a Jacobian integration method [19].

iQ-MS presents volumetric data for individual patients as normalized values; and as centiles referenced to a hypothetical age-matched healthy control. iQ-MS additionally reports brain volumetrics and MS lesion volumes benchmarked to a hypothetical person with MS of similar age, disease duration and disability, to provide a more clinically meaningful, experiential reference. Reference cohorts were created using MRI scans from more than 3,000 healthy controls and an independent sample of 839 people with MS, analyzed with the same methods.

## RESULTS

Of 400 unique scan pairs included in the study, three failed iQ-MS processing due to missing slices in the 3D FLAIR sequence (n=2) or unknown technical reasons (n=1) and were excluded from further analysis. The remaining 397 scan pairs were acquired with a mean interval 12 months (range 6-29 months) from 282 unique patients (F:M=198:84) with a disease duration of 13.1 years (range 0.71-41.83 years) and median EDSS was 1.5 (range 0-7.0, n=315) at the time of the study (follow-up) scan. Incidental findings were present (as determined by the radiology report) in 10.6% of study scans (n=397, Table 3). The vast majority of study scans (387/397) were performed on one of three scanners, each located in different MRI centres: GE MR750 3T (GE Healthcare, Milwaukeee, USA) (n=174), Philips Ingenia 3T (Philips Inc, Amsterdam, The Netherlands) (n=159) and Siemens Skyra 3T (SIEMENS Healthineers, Erlangen, Germany) (n=54). 318/397 scan pairs were acquired on the same scanner with a longitudinally stable protocol defined by iQ-MS (supplementary data).

### Lesion Metrics

Total FLAIR lesion volume determined by iQ-MS was automatically converted to a centile against an (independent) MS patient population built into the tool, and compared against a numerical rating scale/centile assigned to categorical variables in the clinical radiology report, as shown in Table 2. Lesion burden, described in the radiology report of 267/397 unique study (follow-up) scans, matched the equivalent iQ-MS centile in 183/267 (68.5%) of scans; of the remaining scans, the iQ-MS lesion burden fell in a higher centile range in 69/84 (82.1%) of cases. There was a high correlation for both mean FLAIR lesion number (iQ-Solutions: 47.8 [SD 39.0], range 0-223; core lab: 56.0 [SD 44.7], range 1-269; R^2^=0.96,p<0.001) and volume (iQ-Solutions: 6.4mls [SD 10.3], range 0-66.7; core lab: 7.9mls [SD 11.0], range 0-73.1, R^2^=0.96,p<0.001) as detected by iQ-Solutions and the core reading lab. The FLAIR lesion burden also correlated moderately with NBV generated by both iQ-MS (R^2^=0.31,p<0.001) and the core reading laboratory (R^2^=0.24,p<0.001). Disability, as measured by EDSS, correlated only weakly with cerebral FLAIR lesion burden as determined by both methods (R^2^=0.16,p<0.001 and R^2^=0.15,p<0.001 respectively), though significance of the correlation persisted after correction for age, sex, disease duration and brain volume.

**Table 2.** Clinical radiology report FLAIR lesion burden descriptors.

| Clinical descriptor | Numerical assignment | Final assignment | Equivalent iQMS™ Burden (patient* centile) |
| --- | --- | --- | --- |
| • No cerebral lesions<br>• Single lesion<br>• Very scant<br>• Very mild, scant<br>• Relatively scant, mild, relatively mild, small, few, a few, several | 0<br>1<br>2<br>3<br>4 | Mild | <25% |
| • Mild to moderate<br>• Moderate | 5<br>6 | Moderate | 25-75% |
| • Moderately extensive, significant, moderate to heavy, moderate to marked, numerous<br>• Extensive, Severe | 7<br>8 | Severe | >75% |

\*compared with iQ-MS underlying MS reference dataset

**Table 4.** shows the number of scan pairs in which new/enlarging FLAIR lesions or CELs were identified; and the mean new and enlarging lesion numbers for each of the three analysis methods, and for the expert consensus. In total, case-level discrepant results were found in 53/397 case pairs for the presence of new and enlarging lesions; and in 10/180 cases for the presence of CELs. At the lesion number level, discrepancies were present in 51/397 cases (new lesions), 57/397 cases (enlarging lesions), 13/180 cases (CELs), 74/397 cases (new or enlarging lesions), and 78/397 cases (new or enlarging lesions or CELs) among any of the three analysis methods. The outputs and relevant segmentations of all 78 cases exhibiting any discrepancy were manually reviewed (MB, YB) to develop the expert consensus. Visual analysis of twenty randomly selected case-level discrepant pairs (and their analysis outputs) by an independent expert neuro-radiologist (DB), blinded to the expert consensus, corroborated the results of the expert consensus in all cases.

**Table 3.** Distinct incidental findings on MS clinical radiology reports.

| **Incidental Finding** | **Number of Scans (%)** |
| --- | --- |
| Meningioma | 8 (2) |
| Stroke | 4 (1) |
| Focal gliosis | 4 (1) |
| Cavernoma | 3 (0.8) |
| Vestibular schwannoma | 2 (0.5) |
| Nasolabial cyst | 2 (0.5) |
| Intrasellar lesion | 2 (0.5) |
| Developmental venous anomaly | 2 (0.5) |
| Arteriovenous malformation | 2 (0.5) |
| Pineal cyst | 1 (0.3) |
| Neuroglial cyst | 1 (0.3) |
| Left parotid lesion | 1 (0.3) |
| Focal hyperostosis | 1 (0.3) |
| Dural calcification | 1 (0.3) |
| Dilated perivascular space | 1 (0.3) |
| Cryptococcoma | 1 (0.3) |
| Cortical dysplasia | 1 (0.3) |
| Choroid plexus cyst | 1 (0.3) |
| Cerebral artery aneurysm | 1 (0.3) |
| Cerebellar ectopia | 1 (0.3) |
| Cerebral contusion | 1 (0.3) |
| Arachnoid cyst | 1 (0.3) |
| **Total** | **42 (10.6)** |

**Table 4.**
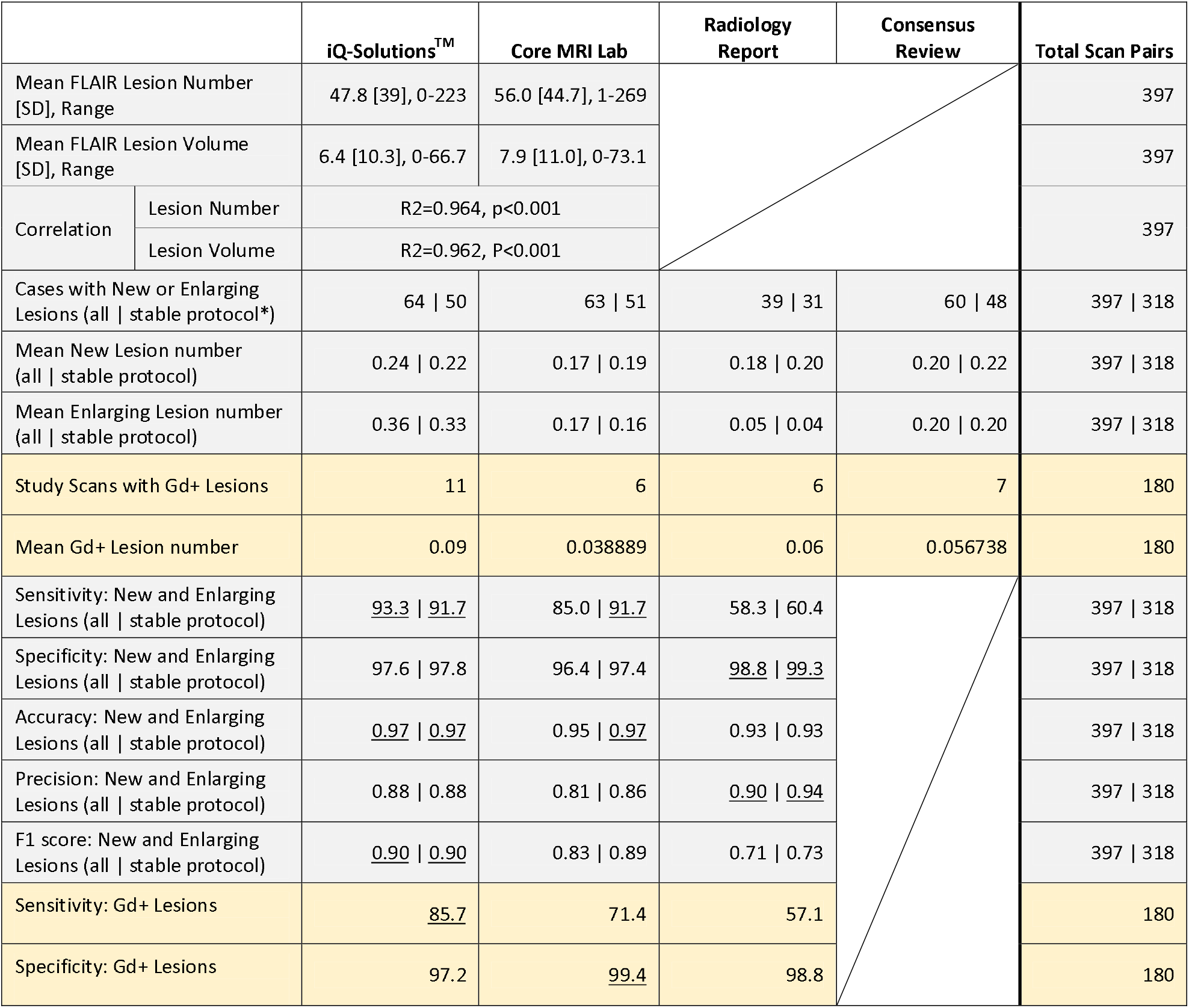
Comparison of Lesion Metrics.

Using the expert consensus as ground truth, iQ-MS more sensitively detected new or enlarging FLAIR (93.3%) lesions and T1-w CELs (85.7%) than either the radiology report (58.3% and 57.1% respectively) or core MRI reading lab (85.0% and 71.4% respectively). When the analysis was restricted to scan pairs with a longitudinally stable scanner/protocol (see supplementary data, n=318), iQ-Solutions and the core MRI reading centre detected new/enlarging lesions with equivalent sensitivity (91.7%), and there was a modest improvement in the sensitivity of the radiology report (60%). Specificity for the detection of FLAIR new/enlarging and T1-w CELs was high for iQ-MS (97.6%, 97.1% respectively), the radiology report (98.8%, 98.8% respectively) and the core MRI reading lab (96.4%, 99.4%); and improved even further when analysis was restricted to scan pairs with a longitudinally stable scanner/protocol (Table 4). For a subset of scans reported by fellowship-trained neuroradiologists (n=268), iQ-MS, radiology reports and the core lab detected MS disease activity in longitudinally stable scans with a sensitivity of 91.0%, 76.0% and 87.9%.

At the lesion level, iQ-Solutions failed to detect an average of 0.02 new lesions per scan using the expert consensus as the gold standard, whereas the core lab and radiology reports failed to detect an average of 0.05 and 0.07 new lesions per scan, respectively. For enlarging lesions, the average number of missed lesions per scan for the three techniques was 0.02, 0.09 and 0.16 respectively.

### Brain Volumetrics

Of the 397 cross-sectional study scans analyzed for brain volume by iQ-MS, 36 cases failed quality control imposed by the core reading lab’s SOP (supplementary data) and were deemed unsuitable for analysis by SIENAX. Comparisons between the methods were therefore restricted to remaining 361 cases. Mean cross sectional brain volume, reported by iQ-MS and the core MRI reading lab (using SIENAX) are shown in Table 5, together with relevant healthy control centile data. NBV was considered to be within normal limits at or above the healthy control 25^th^ centile for both iQ-MS and SIENAX. NBV below this cut-off were identified by these tools in 54.3% and 74.9% of scans respectively; and more severe brain volume loss (≤10^th^ centile) was identified in 32.5% and 38.5% of patients respectively. NBV derived from SIENAX exhibited a greater degree of variance than iQ-MS. Despite these differences, there was a good correlation between NBV derived by the two tools (R^2^=0.671,p<0.001); and NBV correlated, albeit relatively weakly, with EDSS for both (iQ-MS R^2^=0.23,p<0.001; SIENAX R^2^=0.14,p<0.001). Similar observations were made for normalized grey matter and thalamic volumes measured by both iQ-MS and the core MRI reading lab’s implementation of FIRST (Table 5). In a univariate general linear model including NBV, normalized thalamic volume, lesion volume and sex, only NBV (p<0.001) and normalized thalamic volume (p=0.012) were significant contributors to the overall model’s power to predict EDSS (R^2^=0.28, p<0.001). However, the addition of age substantially improved the model’s power (R^2^=0.35, p<0.001) and rendered the contribution of NBV non-significant (p=0.352), while preserving the significance of normalized thalamic volume (p<0.001) as a significant predictor of EDSS, in keeping with the known association of this structure with MS disease progression [20]. An analogous pattern was observed using metrics derived from the core lab. Notably, radiology reports only described the presence or absence of brain volume loss in 99/397 study scans, of which 23% were reported to have some degree of brain volume loss, though this was not categorized in vast majority, preventing meaningful statistical comparison. None of the radiology reports described thalamic volume change.

**Table 5.** Comparison of Brain Volumetrics.

| Cross-sectional volumetrics, n=361 unique scan pairs |  |  |  |  |
| --- | --- | --- | --- | --- |
|  |  | iQ-Solutions™ | Core MRI Lab^ | Radiology Report |
| Mean Normalized* Brain Volume [SD] (mls) |  | 1460.4 [56.2] | 1427.4 [77.0] |  |
| Mean Normalized Thalamic Volume [SD] (mls) |  | 18.2 [2.1] | 19.2 [2.2] |  |
| Mean Normalized Grey Matter Volume [SD] (mls) |  | 772.3 [38.3] | 755.5 [48.9] |  |
| NBV HC Centile® (%) | ≥25<sup>th</sup> | 44.9 | 13.0 | 19.1 ('No BVL') |
|  | 10<sup>th</sup>-25<sup>th</sup> | 20.2 | 27.4 | 5.8 ('BVL') |
|  | ≤10<sup>th</sup> | 34.9 | 59.6 | 75.1 (Unknown) |
| Normalized Brain Volume Correlation |  | R<sup>2</sup>=0.67,p<0.001 |  |  |
| Normalized Thalamic Volume Correlation |  | R<sup>2</sup>=0.80,p<0.001 |  |  |
| Normalized Grey Matter Volume Correlation |  | R<sup>2</sup>=0.69,p<0.001 |  |  |
| Longitudinal volumetrics, n=295 unique scan pairs |  |  |  |  |
|  |  | iQ-Solutions™ | Core MRI Lab^ | Radiology Report |
| Mean Interval PBVC Change [SD] (%) |  | -0.32 [-0.73] | -0.36 [-0.71] |  |
| PBVC Correlation |  | R<sup>2</sup>=0.86, p<0.001 |  |  |
| Interval PBVC Severity (Loss) n (%) | <0.4% | 161 (54.6%) | 161 (54.6%) | 80% ('No atrophy') |
|  | 0.4-0.8% | 70 (23.7%) | 70 (23.7%) | 20% (Unknown) |
|  | >0.8% | 64 (21.7%) | 64 (21.7%) |  |
| Interval Thalamic Volume Change [SD] (%) |  | -0.33 [1.75] |  |  |
| Interval Grey Matter Volume Change [SD] (%) |  | -0.15 [1.02] |  |  |

NBV: normalized brain volume, PBVC: percent brain volume change.

Mean interval brain atrophy was calculated for all pairs that passed longitudinal analysis criteria defined by the iQ-Solutions automated protocol QC/analysis (n=318). Of these pairs, a further 23 failed quality control imposed by the core reading lab’s relevant SOP (supplementary data) and were deemed unsuitable for analysis by SIENA. Comparisons between the methods were therefore restricted to remaining 295 scan pairs. Mean annualized PBVC was similar for both methods (iQ-MS: -0.32% [SD -0.73%]; SIENA: -0.36% [SD -.71%]). There was a strong correlation (R^2^=0.86, p<0.001) between annualized PBVC determined by the two methods (Figure 2). Using a pathological cut-off of 0.4% PBVC [21], brain atrophy was detected in 134/295 (45.4%) of study scans using both quantitative methods; and severe brain atrophy (>0.8% per year) was also equivalently detected in 64/295 (21.7%) of scans. However, at the individual scan level, classification of annualized atrophy as severe (>0.8%) was discordant in 24/295 cases. Of these cases, a difference of more than 0.2% was observed in 15/24 between iQ-Solutions (greater atrophy in 8 patients) and SIENA (greater atrophy in 7 patients). Qualitative assessment of brain volume change in scan pairs was described in 236/295 radiology reports; no interval atrophy was reported in any of the assessed scan pairs.

**Figure 2.**
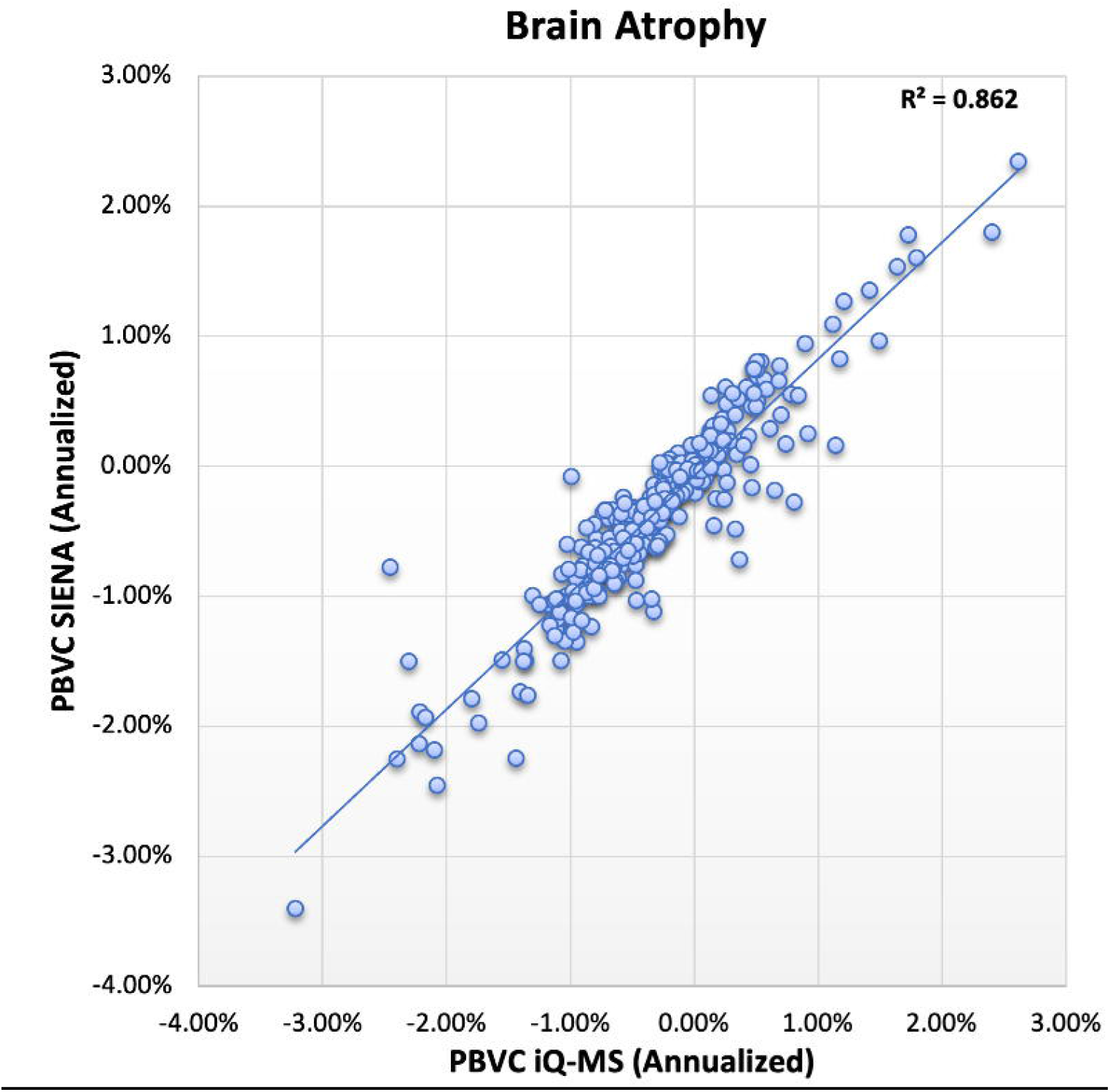
Correlation of annualized percentage brain volume change between scan pairs as determined by SIENA and iQ-MS.

Interval iQ-MS PBVC was weakly correlated with both new (R=-0.11,p<0.05) and enlarging (R=-0.13,p<0.01) lesion volume; and survived partial correlation correction for age, sex and disease duration. SIENA-derived PBVC was not correlated with any of these variables. There was also a weak correlation of PBVC, as derived by both methods, with EDSS that survived correction for age, sex and disease duration (Table 6).

**Table 6.** Correlations with Disability Status.

| Clinical Correlations (disability status at study scan) |  |  |
| --- | --- | --- |
|  | iQ-Solutions™ | Core MRI Lab^ |
| **NBV-EDSS** | $R^2=0.23, p<0.001$ | $R^2=0.14, p<0.001$ |
| **NTV-EDSS** | $R^2=0.21, p<0.001$ | $R^2=0.17, p<0.001$ |
| **GMV-EDSS** | $R^2=0.23, p<0.001$ | $R^2=0.12, p<0.001$ |
| **PBVC-EDSS** | $R^2=0.05, p<0.001$ | $R^2=0.03, p=0.04$ |
| **Lesion Volume-EDSS** | $R^2=0.16, p<0.001$ | $R^2=0.15, p<0.001$ |
| **New Lesion Volume-EDSS** | NS |  |
| **Enlarging Lesion Volume-EDSS** | $R^2=0.05, p<0.001$ | |

NBV: normalized brain volume, NTV: normalized thalamic volume, GMV: normalized grey matter volume, PBVC: annualized percent brain volume change, EDSS: expanded disability status scale

## DISCUSSION

We demonstrate superior performance of the fully automatic, deep learning based tool, iQ-MS, for detection of new, enlarging and contrast-enhancing lesions, the principal indicators of subclinical MS disease activity, compared with qualitative radiology reporting. We also show at least equivalent performance of the AI tool with semi-automated quantitative lesion activity and volumetric assessments undertaken by an experienced, ISO-9001 certified core imaging laboratory.

The detection of clinically silent new MRI lesions is an important determinant of treatment strategy [4] that may, in its own right, lead to escalation of immunotherapy. Most modern therapeutic paradigms target No Evidence of Disease Activity (NEDA) [7], which encompasses both clinical and radiological disease quiescence. Here, we report a case-level sensitivity of 93.3%, relative to a consensus ground truth, for detecting MS disease activity in multi-centre, real-world MRI scans acquired approximately 12 months apart, an interval consistent with recommended routine clinical practice for monitoring MS and treatment efficacy. For this metric, the fully automatic AI based tool substantially outperformed clinical radiology reports (sensitivity 58.3%), despite only a minor sacrifice in specificity (97.6% vs 98.8%); and was at least equivalent to the core imaging laboratory (sensitivity 91.7% in longitudinally consistent scans for both methods). Not surprisingly, when a subset of scans reported by fellowship-trained neuroradiologists was analyzed, the sensitivity of radiology reports for MS disease activity rose substantially (77%), but remained essentially stable at 92.3% for iQ-MS (data not shown).

At the lesion level, iQ-Solutions missed the equivalent of only 1 new lesion for every 44 scans analyzed in our cohort, whereas conventional radiology reports and the core lab missed the equivalent of 1 new lesion in 15 and 19 scans respectively. Using the average new lesion volume calculated by the AI tool and an approximation of the number of axons transected per mm^3^ of new lesional tissue [22], the application of iQ-MS therefore represents a potential opportunity to prevent, with appropriate treatment change, an averaged irreversible loss of >45,000 (up to >2 million in individual patients) axons, over 12 months relative to individuals monitored with conventional radiology reporting alone. These numerical extrapolations assume the availability of a therapy that can effectively prevent new lesion formation.

However, improved sensitivity for interval disease activity relative to radiology reports was driven primarily by failure of the human reporter to capture enlarging lesions consistently, perhaps not an unexpected finding given their visual subtlety in comparison with new, free-standing lesions. Enlarging lesions are under increasing scrutiny as a primary driver of disability worsening, especially for patients in whom relapses have been essentially abolished by high efficacy DMT. In particular, slowly enlarging lesions (SELs), likely an imaging surrogate of ‘smouldering’ MS lesions that exhibit chronic inflammation at their edge [23], have gained traction as an independent biomarker of disease progression with a distinct pathophysiology [24]. Currently, iQ-MS does not isolate concentrically enlarging lesions from the global enlarging lesion pool, nor does it automatically monitor individual lesions over multiple timepoints to separate subacute from slow lesion enlargement. Incorporating these capabilities into AI-based lesion activity tools such as iQ-MS will become more pressing as pharmacotherapies that putatively target these pathomechanisms, such as the BTKi drugs, are developed [25].

The detection of contrast enhancement, a marker of blood-brain barrier disruption that characterises new MS lesion formation and typically persists for 2-6 weeks, was also assessed in the 180 study scans in which gadolinium contrast was administered. While the sensitivity of iQ-MS for this metric (85.7%) was less impressive than for new and enlarging lesions, the fully automatic AI-based tool significantly outperformed (Table 4) the other methods with only a minor impact on specificity (97.2%). The omission of gadolinium administration from routine MS monitoring protocols [26,27] further emphasises the need for tools that sensitively detect interval development of new and enlarging lesions.

Accelerated brain atrophy occurs at the earliest stages of MS and is a recognised marker of neurodegeneration [28]. The role of brain volumetrics in the clinical management of individual people with MS is less well defined. At the group level, there is good evidence that lower cross-sectional normalized brain volumes correlate with worse disability outcomes [29]; and that short term (1-2 years) brain atrophy can predict longer term clinical outcomes [30]. Translation to individual patients is confounded by measurement error inherent to analysis techniques; longitudinal scan acquisition inconsistency; and biological and treatment-related fluctuations in brain volume [31]. However, brain volume below the 10^th^ centile of an age-matched healthy control, a consistent adverse brain atrophy trajectory over multiple clinical epochs or severe PBVC (>0.8% per annum) over a single epoch, may influence or support changes in immunotherapy in conjunction with relevant clinical and lesion metrics. The current literature lacks clinical evaluation and validation data for existing quantitative volumetric reports for people with MS [14].

Here, we report high correlations between the outputs of the AI tool and both cross-sectional and longitudinal whole brain volumetric tools measured in a core MRI lab using SIENAX [32] (R^2^=0.67) and SIENA [32] (R^2^=0.86) respectively; and comparatively improved correlations with the EDSS (Table 6). Severe brain volume loss (<10^th^ healthy control centile) was present in substantial proportion of study scans, but cross-sectional brain volume loss of any severity was only mentioned in a small proportion (<25%) of radiology reports, potentially reflecting assumed lack of clinical relevance of this metric by the reporting radiologist or inability of the human reporter to assess brain volume loss, even qualitatively, relative to a hypothetical, age-matched healthy control. The principal measure of clinical interest, annualized PBVC, was similar across the two quantitative tools (iQ-MS mean PBVC -0.32%, SIENA mean PBVC -0.36%), and fell within the range (<0.4% loss) considered non-pathological [21]. This is unsurprising in a modern MS cohort, given that many of the highly effective therapies, use of which is prevalent in Australia, ameliorate brain volume loss in randomised clinical trials [33-35]. When stratified by severity, the tools appear to show equivalent interval PBVC among the atrophy subgroups when referenced to the same healthy control cohort analyzed with the respective methods (Table 5). However, the presence of severe atrophy (>0.8% negative PBVC), as determined by the two quantitative methods, was discordant in 24/295 (8.1%) of cases, highlighting methodological concerns when applying these tools to individual patients over single, relatively short epochs. Compared to SIENA, we have recently shown that DeepBVC, the brain atrophy algorithm embedded in iQ-MS, demonstrates greater stability and superior performance in test-retest experiments; and is more robust to variance in imaging acquisition [18]. Likely reflecting inability of the human reporter to detect minor brain volume changes over short intervals, no interval atrophy was reported in any of the 236/295 scan pairs that were visually assessed.

When a clinician evaluates brain imaging in a person with MS, they mentally compare the scan before them not only with a hypothetical healthy person of similar age, but also a hypothetical patient, derived from their cumulative experience, of similar age, disease duration and treatment. In conventional monitoring paradigms, such a comparison is necessarily indirect, qualitative and limited by the experience of the reporting radiologist and clinician. Existing quantitative imaging tools partly address this through comparison of individual patients to healthy controls, as does the fully automatic, AI tool described here. To our knowledge, this is the first tool to additionally embed an ‘experiential’ comparator that returns a relevant patient centile for both FLAIR lesion burden and brain volumetric data. While the clinical utility of this additional information is unknown, the integration of iQ-MS into the recently inaugurated MSBase Imaging Repository will facilitate the development of a broader comparative experiential dataset that can be properly benchmarked in research settings.

Finally, incidental findings (Table 3) were reported by the radiologist in 10.6% of MS scans. We emphasize that iQ-Solutions™ is a non-diagnostic tool designed for the quantitative monitoring of people with a known neurological disease, here applied to MS, to facilitate their precision treatment. Radiologist oversight, both for quality control of the results provided by the AI tool; and for reporting clinically significant incidental findings, remains paramount.

Our study has a number of limitations. The bulk of MRI scans in the study were acquired on one of 3 scanners, potentially limiting the generalizability of the results. Additionally, most scan pairs (318/397) analyzed in our cohort were acquired on the same scanner with a consistent imaging protocol, as determined by iQ-MS, that may be difficult to enforce in some clinical settings. Although not a principal outcome of our study, the high correlation between cross-sectional lesion number/volume as determined by iQ-MS and the core lab should be interpreted with caution, given that image analysts in the core lab manually adjusted lesion masks that were initially created with an in-house AI algorithm that shared training data with the fully automatic solution. However, longitudinal lesion metrics (new and enlarging lesions), the outcome of most clinical relevance, were manually determined by the core lab via the aid of an independent subtraction image and slice by slice visual inspection. While the determination of the expert consensus was potentially confounded by lack of blinding (imposed by the distinct formats of the segmentations reviewed by the expert neurologist and neuroradiologist), the consensus was corroborated by an independent radiologist in all case-level discrepant scan pairs reviewed. Volumetric performance of the AI tool was confined to scan pairs with an available quantitative comparator. However, normalized brain volume and PBVC correlated strongly with de-facto gold standards used in the majority of modern MS clinical trials, exhibited less variance than these comparators, and better, though still weakly, correlated with a measure of clinical disability.

## Conclusions

iQ-MS is a sensitive and accurate tool for monitoring MRI scans in people with MS by providing quantitative metrics that value-add to traditional radiology reports. Comparison with both radiology reports and a core MRI analysis lab shows superiority of the AI tool across a range of lesion and volume measures derived from clinically acquired, multicentre scans. The incorporation of a novel, experiential reference provides a more clinically meaningful quantitative comparator for lesion burden and brain volumetric analyses. The scaled deployment of AI-based quantitative imaging tools, such as iQ-MS, has the potential to enhance both real-world, clinical-imaging disease-specific research and the precision management of individual patients with MS.

## METHODS

### Patients and Clinical Data

Patients with a diagnosis of MS attending the Royal Prince Alfred Hospital MS Service were retrospectively included in the study. The study was approved by The University of Sydney Human Research Ethics Committee and followed the tenets of the Declaration of Helsinki. Written informed consent was obtained from all participants. De-identified clinical data, including diagnosis, disease duration (from symptom onset), gender, age in years and expanded disability status scale (EDSS) score, were extracted through the clinic’s MSBase [36] interface.

### Imaging Data and Informatics

Inclusion criteria included a minimum of two available MRI timepoints, separated by at least 6 months. Scans with 3D T1-w and 3D FLAIR imaging, acquired on any MRI scanner, were included in the study; there were no pre-specified sequence parameters. Based on a significance level of 5%, assumed 80% sensitivity of radiologist reports for detection of MS lesion activity, and power of 80% to identify a 10% improvement with iQ-MS, recruitment to the study ended when 400 appropriate scan pairs had been included. All images were automatically de-identified with an informatics tool, Torana™ (Sydney Neuroimaging Analysis Centre, Sydney), prior to their inclusion in an in-house research PACS for automatic analysis and processing by iQ-MS. The MSBase identifier was automatically added to the image meta-data to facilitate subsequent matching with the patient’s clinical data. To simulate real-time clinical workflow, annotations and reports generated by iQ-Solutions were automatically returned to Torana™ and transferred into the appropriate project/subject/scan session in the PACS for review by study staff.

### Clinical Radiology Reports

Clinical radiology reports were de-identified and then reviewed by an expert MS neurologist (MB), who extracted and recorded the following metrics: number of new FLAIR lesions, enlarging FLAIR lesions and T1-w CELs. Scans were categorized as active if any new/enlarging or enhancing lesions were detected. The burden of cerebral FLAIR MS pathology, where reported, was recorded and an attempt made to transform descriptors into a numerical rating made (Table 2). The presence/absence of reported brain volume loss and its severity (mild, moderate, severe) was recorded, as was the presence/absence of brain atrophy between current and prior studies. Incidental findings and their type were recorded. Reports were also categorized by whether the reporting radiologist was a fellowship-trained subspecialty neuro-radiologist or a general radiologist.

### Core MRI Reading Laboratory

All scans were independently (and blindly) analyzed by trained neuroimaging analysis staff at the Sydney Neuroimaging Analysis Centre, an ISO-9001 certified core MRI reading facility, using standard operating procedures (SOP) designed for regulatory MS clinical trials. FLAIR lesion number and volume was iteratively measured on intensity-inhomogeneity corrected 3D FLAIR imaging using an in-house lesion segmentation tool, followed by manual quality control of every image slice and lesion mask adjustment with a semi-automated thresholding technique. Importantly, the in-house tool for measurement of these (cross-sectional) lesion metrics used an AI algorithm that shared training data with the fully automatic solution used by iQ-MS. Lesion activity analysis (the development of new or enlarging lesions) was performed manually with the aid of a subtraction image and slice by slice inspection. Enlarging lesions were defined as any pre-existing FLAIR hyperintensity that had enlarged, either concentrically or eccentrically, between the prior and current scan on ≥2 consecutive slices. Scans were categorized as active if any new/enlarging or enhancing lesions were detected. CELs were identified on co-registered post contrast 3DT1 images and enhancing voxels segmented using a semi-automated thresholding technique.

Quality control determined by the core lab’s SOP for observational studies was implemented to exclude scans unsuitable for cross sectional or longitudinal analysis (supplementary data). At each time point, the quantification of absolute and normalized brain volume (NBV) and thalamus volume was estimated on co-registered pre-contrast lesion in-painted and inhomogeneity corrected 3D T1-w images using FMRIB’s SIENAX (version 2.6) [32] and FIRST [37] software packages respectively. Quantification of longitudinal percentage brain volume change (PBVC) between the current and prior scan was determined by a modified hybrid of FMRIB’s SIENA [32] software. Annualized brain atrophy was categorized as normal (<0.4%), mild-moderate (0.4-0.8%) or severe (>0.8%).

### iQ-Solutions™ MS Report

All iQ-MS data was derived automatically using the workflow described under Imaging Data and Informatics above; for clarity, no human intervention was introduced at any point. Specific metrics returned by iQ-MS are shown in Table 1. FLAIR MS lesion burden was categorized using the automatically returned patient (MS population) centile figure (see Introduction, Table 2) as mild (<25^th^ patient centile), moderate (25^th^-75^th^ patient centile) or severe (>75^th^ patient centile). Brain volume loss was categorized using the automatically returned HC centile figure (Table 2) as none (≥25^th^ HC centile), mild-moderate (10-25^th^ HC centile) or severe (≤10^th^ HC centile). Annualized brain atrophy was categorized as normal (≤0.4%), mild-moderate (>0.4%,≤0.8%) or severe (>0.8%), based on previously determined ‘pathological cut-offs’ of brain atrophy using SIENA [21], against which the relevant iQ-MS algorithm has been previously validated [18].

### Expert Consensus

To establish a ground truth, a case and lesion level comparison of the output of each method (radiology report, neuroimaging analyst, iQ-MS) was undertaken. Where there was agreement at both the case (active vs inactive) and lesion (number of new lesions, enlarging lesions and CELs) level across all three methods, the results were accepted as the ground truth. Where any discrepancy was noted at either the case or the lesion level, further review of the raw images, together with the output of all three methods (including final segmentations from both neuroimaging analysts and iQ-MS) was undertaken by an expert neuro-radiologist (YB) and neurologist (MB) and a final ground truth established by consensus. As the segmentation masks output by the three methods differed in both format and visual appearance, this review was necessarily unblinded. As such, a random sample of >25% of all case-level discrepant scan results was reviewed by a third, independent expert neuroradiologist to determine conformity with the expert consensus.

### Statistics

Statistical analyses were performed using SPSS 26.0 (SPSS, Chicago, IL, USA). Descriptive statistics were calculated for all inter-method comparisons. Subgroups studied included (i) scans reported by a subspecialist neuroradiologist and (ii) scan pairs acquired with a longitudinally consistent imaging protocol as defined in supplementary data. Pearson’s correlation coefficient was used to measure statistical dependence between two numerical arrays, and p< 0.05 was considered statistically significant. For partial correlation, data were adjusted for age, gender and disease duration; p⍰< ⍰0.05 was considered statistically significant.

## Data Availability

The datasets analysed in the current study are available from the corresponding author on reasonable request and with a relevant research agreement.

## DATA AVAILABILITY

The datasets analysed in the current study are available from the corresponding author on reasonable request and with a relevant research agreement. The underlying code for iQ-Solutions™ is not publicly available for proprietary reasons; however, code for specific individual algorithms is described in the relevant references [16-18].

## SUPPLEMENTARY DATA

### Core lab quality criteria for assessment of scan suitability for cross sectional analysis brain volumetric analysis

The core lab standard operating procedure (SOP) for observational studies was implemented in the current work. Images were excluded from cross-sectional analysis volumetric analysis if:

1. Acquisition problems, based on visual inspection, are present in scan, namely:
  a. the brain was not scanned from the base of the brain stem to the vertex.
  b. inadequate tissue contrast, spatial contrast or image inhomogeneity that is likely to compromise analysis quality
  c. images contain artifacts such as aliasing, Gibbs or Truncation, Zipper, Motion or Susceptibility artifacts that are likely to compromise analysis quality
2. Image slice thickness, image geometry and detailed parameter settings are study-specific and specified by the core lab external to the SOP. For the current study, gapless 3DT1 and 3DFLAIR (for lesion in-painting) sequences with a thickness of ≤3mm were specified, but no specific parameter settings were pre-specified.

### Determination of longitudinal scanner/protocol stability for brain volumetric analysis

#### iQ-MS

Longitudinal inconsistency between timepoints is determined by any of:

1. Scanner mismatch: scans are acquired on a different scanner at the two timepoints
2. Protocol mismatch: scans at the two timepoints are not acquired with similar protocols. Similar protocols are defined as protocols in which the Acquisition Voxel Size does not change by more than 30%. The Acquisition Voxel Size is calculated by [Row Pixel Spacing] * [Column Pixel Spacing] * [Number of Rows] * [Number of Columns] * [Slice Thickness] * [Non-zero elements in the Acquisition Matrix] where all referenced quantities are extracted from the DICOM headers.
3. Affine similarity mismatch: scans at the two timepoints have affine similarity <0.2, implemented as described previously [1]

#### Core Lab

The core lab standard operating procedure (SOP) for observational studies was implemented in the current work. Relevant elements of the SOP are included below.

Longitudinal inconsistency between timepoints is determined by any of:

1. Scanner mismatch: scans are acquired on a different scanner at the two timepoints
2. Acquisition problems, based on visual inspection, are present in scans at either timepoint, namely:
  a. the brain was not scanned from the base of the brain stem to the vertex.
  b. inadequate tissue contrast, spatial contrast or image inhomogeneity that is likely to compromise analysis quality
  c. images contain artifacts such as aliasing, Gibbs or Truncation, Zipper, Motion or Susceptibility artifacts that are likely to compromise analysis quality
3. Protocol (image geometry and detailed parameter settings) inconsistency: these metrics are study-specific and specified by the core lab external to the SOP. For the current study, no specific parameter deviations were pre-specified.

