## SUPPLEMENTARY DATA for "AI-BASED MRI MONITORING IN MULTIPLE SCLEROSIS: REAL-WORLD CLINICAL VALIDATION"

(d) the brain was not scanned from the base of the brain stem to the vertex.
(e) inadequate tissue contrast, spatial contrast or image inhomogeneity that is likely to compromise analysis quality
(f) images contain artifacts such as aliasing, Gibbs or Truncation, Zipper, Motion or Susceptibility artifacts that are likely to compromise analysis quality

3. Protocol (image geometry and detailed parameter settings) inconsistency: these metrics are study-specific and specified by the core lab external to the SOP. For the current study, no specific parameter deviations were pre-specified.

**Supplementary Data References**

1. Sima D. M., Horáková D., Nguyen A.-L., Van Hecke W., Kalincik T., Barnett M. H., et al. (2019). Assessing the reliability of longitudinal MRI examinations in multiple sclerosis follow-up. ECTRIMS Online Libr. 278907:547.
